# Real-time Volumetric Imaging for Minimally-Invasive Thoracic Interventions: A Deformable Phantom Study

**DOI:** 10.64898/2026.10.28.26366767

**Authors:** Alexander Lin, Alen Biju, Allen Goode, Jessica Im, Peter Noel, Magdalena Bazalova-Carter, Tess Reynolds, Nicholas Hindley

## Abstract

**Objectives:** This study aimed to evaluate the ability of VoxelMap, a deep learning framework for 3D respiratory motion estimation, to generate volumetric images in real-time from 2D projections acquired for two deformable lung phantoms on a clinical interventional C-arm angiography system.

**Materials and Methods:** VoxelMap was evaluated using the PixelPrint^4D^ and Casper deformable physical thoracic respiratory phantoms. Four-dimensional cone-beam CT (4D CBCT) images acquired on a clinical fixed-room C-arm angiography system were used for network training and volumetric ground truth. Prediction accuracy was evaluated on withheld respiratory phases using structural similarity index measure (SSIM) and peak signal-to-noise ratio (PSNR). Model inference time was measured to assess suitability for continuous real-time processing. Deep ensembles were also used to assess model uncertainty. Correlation between ensemble uncertainty and prediction error was evaluated for both phantoms.

**Results:** VoxelMap achieved high prediction accuracy across withheld respiratory phases in both physical phantoms. SSIM values exceeded 0.92 in all experiments, with a higher performance observed for the Casper phantom (0.969-0.973) than for the PixelPrint^4D^ phantom (0.927-0.943). Model inference times were less than 15 ms for both phantoms which is compatible with real-time processing. Ensemble uncertainty was positively correlated with prediction error in both phantoms, indicating that higher uncertainty was associated with less accurate volumetric predictions.

**Conclusions:** Our study demonstrated that VoxelMap generates accurate volumetric deformations of respiratory anatomy from projections acquired on a clinical interventional C-arm system, with computational inference times compatible with continuous processing. By deriving volumetric information directly from two-dimensional fluoroscopic projections, VoxelMap has the potential to augment continuous fluoroscopic guidance with continuously updated volumetric information. Ensemble uncertainty may provide a potentially useful indicator of prediction reliability. Further studies using clinically representative training data and patient studies is required to establish clinical utility.

Our code is available at https://github.com/Image-X-Institute/Voxelmap.

## 1 Introduction

Many image-guided interventions rely on X-ray fluoroscopy to provide continuous visualization during procedures [1, 2, 3]. 2D fluoroscopy is well suited for interventions given its excellent temporal resolution, ready availability, low acquisition burden, and direct integration into many interventional procedures [4, 2, 3]. However, all 2D techniques infer rather than directly visualize the underlying 3D anatomy [5]. This limits the navigational utility of fluoroscopy across several applications including catheter navigation, needle placement, tumor targeting, endovascular interventions, device deployment and structural procedures [1, 3, 6].

Intra-procedural 3D cone beam computed tomography (CBCT) can provide volumetric anatomical information and therefore has become an important adjunct to 2D fluoroscopic guidance [6]. Specifically, CBCT imaging in thoracic intervention supports procedural navigation and confirmation of probe placement, increasing navigational success and diagnostic yield [7, 6, 8, 9]. Applications include diagnosis of peripheral pulmonary lesions, CBCT-guided navigation bronchoscopy, navigating biopsy devices, and CBCT-guided video assisted thoracic surgery [8, 10, 11, 12]. However, CBCT and all pre-intervention 3D scans necessarily represent outdated information rather than continuous updates of the current state of anatomy [13]. This is clinically relevant in thoracic interventions where the relationship between the underlying pulmonary anatomy, lesions, and instruments changes with respiration [14]. Furthermore, dynamic anatomical changes including atelectasis and bronchoscope-induced airway deformation can obscure target lesions on intra-procedural CBCT, reducing the diagnostic yield of CBCT-guided bronchoscopy [15, 16].

Digital tomosynthesis (DTS) attempts to provide depth information with a lower acquisition burden, using a limited angle acquisition with fewer projections to produce a quasi-3D reconstruction [17]. However, the limited angle acquisition results in poor depth resolution which limits the role of DTS in providing reliable 3D localisation for thoracic interventions [18, 17]. Neither CBCT nor DTS provide continuously updated volumetric information throughout an intervention.

Currently, there is no continuously updated volumetric guidance derived from 2D fluoroscopic images. One approach to this problem is VoxelMap, a deep learning framework for real-time volumetric imaging during image guided interventions. VoxelMap trains a neural network to learn patient-specific correspondences between intra-procedural 2D imaging and the underlying 3D anatomy. Originally developed in the context of image-guided radiotherapy, this framework enables the generation of continuous real-time volumetric reconstructions by using existing fluoroscopic imaging to deform a reference volumetric image. VoxelMap has demonstrated the production of volumetric deformations with a high similarity to ground truth on several physical and synthetic phantoms, demonstrating algorithmic feasibility for reconstructing respiratory motion [19, 20]. The next translational step is demonstrating that VoxelMap achieves accurate volumetric deformations using projections acquired on clinical interventional systems and physical respiratory phantoms.

The application of artificial intelligence (AI) in an interventional setting raises questions regarding the reliability and safety of model predictions. As deep learning networks are prone to unpredictable failures, transparency and explainability in model development and validation are critical to establish clinician trust and support safe clinical adoption [21, 22]. Clinicians have shared that explainability involves models highlighting where their predictions may be unreliable; models that provide transparency around their limitations may remain acceptable despite underperformance [23]. Clinician mistrust is a key barrier to the adoption of deep learning in healthcare and uncertainty estimation has been proposed as a mechanism to identify potentially inaccurate predictions and support clinical trust [24]. An uncertainty estimate that is correlated with prediction error could indicate when model predictions should be interpreted with greater caution. Deep ensembles are an established method to estimate the uncertainty of model prediction [25]. Therefore, in this study, we extended the VoxelMap architecture with a deep ensemble network and evaluated the correlation between prediction uncertainty and prediction error. This approach produces both a volumetric prediction and an estimate of its reliability, addressing two key requirements for the interventional application of VoxelMap: continuous volumetric information and confidence in the resulting predictions.

To address the need for continuously updated and reliable volumetric information in interventional settings, the contributions of this paper are:

1. first demonstration of real-time volumetric imaging using 2D projections acquired on a clinical fixed-room C-arm angiography system;
2. evaluation of VoxelMap prediction performance across two motion programmable physical respiratory phantoms and;
3. leveraging deep ensembles to provide an intuitive indicator of model uncertainty.

## 2 Materials and methods

### 2.1 PixelPrint^4D^ phantom

The deep-learning VoxelMap architecture was first trained on images acquired for a physical single-lung phantom (PixelPrint^4D^), shown in Figure 1. The PixelPrint^4D^ phantom was modeled on a radiation oncology patient lung and mimics the structures of the reference lung including the bronchi, vessels, tumor, lung fissures, and heterogeneities in the lung parenchyma, with a structural similarity index measure (SSIM) of 0.93 [26]. The lung phantom demonstrated similar CT attenuation (Hounsfield unit (HU)) and structural features relative to the reference lung, with the mean absolute error as 50 *±* 64 HU and SSIM as 0.93 over the total lung volume [26]. e phantom was deformed simultaneously by anterior/posterior (AP) and superior/inferior (SI) compressors along the AP and SI directions respectively to mimic the deformation of the reference lung over the respiratory cycle, with level of compression controlled by adjusting attachment screws.

**Figure 1:**
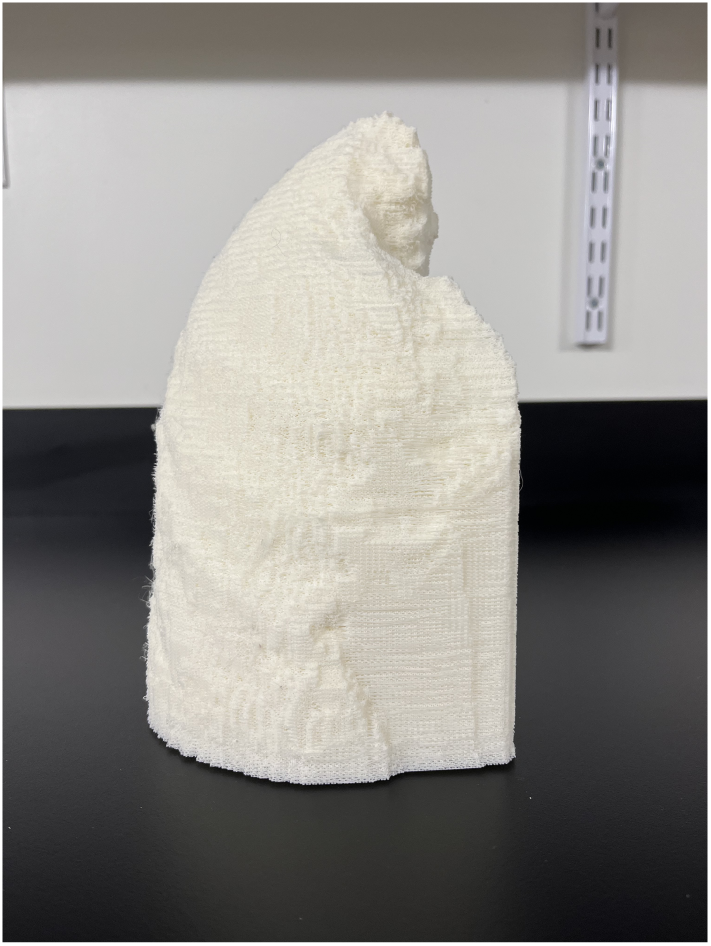
The PixelPrint^4D^ phantom.

Eight breathing phases were defined with phase 1, corresponding to end inhalation, as the reference with diaphragm AP displacement 0 mm and SI displacement 0 mm. The remaining phases were defined as shown in Table 1, with phase 6 (AP 2.2 mm, SI 15.0 mm) corresponding to end exhalation and phase 8 (AP 0.6 mm, SI 3.6 mm) corresponding to mid-inhalation. The deformation of the phantom between phase 1 and phase 6 is similar to that of the reference lung between end inhalation and end exhalation, with mean attenuation changes and percentage volume differences within 5 HU and 4% difference compared to the reference, respectively [26].

**Table 1:** Phase definitions for PixelPrint^4D^ Phantom with Diaphragm Anteroposterior (AP) and Superior-Inferior (SI) Displacements.

| **Phase** | **Description** | **AP Displacement (mm)** | **SI Displacement (mm)** |
|---|---|---|---|
| 1 | End inhalation (Reference) | 0.0 | 0.0 |
| 2 | – | -0.6 | 4.1 |
| 3 | – | -0.5 | 8.9 |
| 4 | – | 0.3 | 12.7 |
| 5 | – | 1.2 | 14.7 |
| 6 | End exhalation | 2.2 | 15.0 |
| 7 | – | 1.7 | 9.2 |
| 8 | Mid-inhalation | 0.6 | 3.6 |

4D CBCT was chosen as the training and ground-truth imaging modality to provide a controlled extension of the previously published VoxelMap framework, which used 4D-CT data [19, 20]. This enabled evaluation of VoxelMap using projections acquired on a clinical interventional C-arm while retaining volumetric ground truth for respiratory motion.

CBCT images were obtained on a fixed-room C-arm angiography system (Siemens ARTIS Pheno, Siemens Healthineers GmbH, Forchheim, Germany) within the Department of Radiology and Medical Imaging of UVA Health University Medical Centre using the 5s DynaCT (DCT) Body protocol at the eight breathing phases. Each 3D scan was generated from 2D X-ray projections at 397 imaging angles over one 200*^◦^* arc to produce a total of 3176 2D projections. The PixelPrint^4D^ images were preprocessed by resizing the 3D images to dimensions 128 x 128 x 128 and the 2D projections to 128 x 128 due to memory constraints.

### 2.2 Casper phantom

Cross-phantom validation was performed on a physical thoracic phantom (Casper), shown in Figure 2. The Casper phantom mimics the human torso, including lungs, ribs, and chest wall [27, 7]. Each component has a similar HU to the corresponding human anatomy including lungs (−840 HU), ribs (125 HU), and soft tissue (46 HU) [27]. Deformations produced by a CIRS Dynamic Thorax motor (Sun Nuclear Corporation) attached to the diaphragm plate, which alters the diaphragm position along the SI axis, are similar to those produced by human respiratory motion [7].

**Figure 2:**
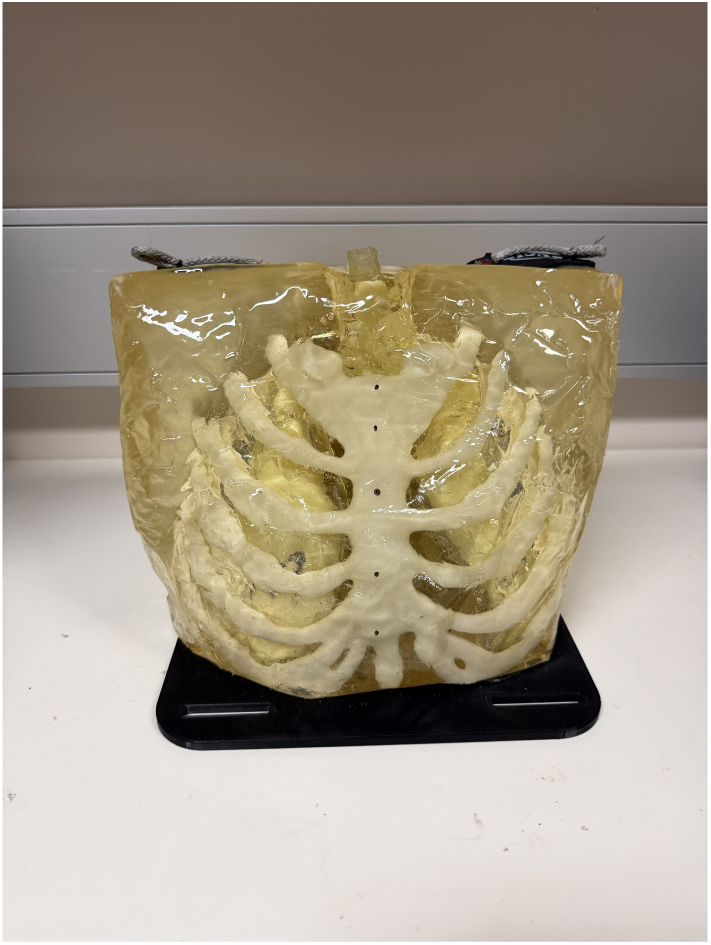
The Casper phantom.

CBCT images were obtained with a fixed-room C-arm angiography system (Siemens ARTIS Pheno, Siemens Healthineers GmbH, Forchheim, Germany) in the Hybrid Theatre, Sydney Imaging, University of Sydney using the 5s DCT Body scan protocol in ten breathing phases between the diaphragm positions −6.5 mm and +7.0 mm relative to the reference diaphragm position. Phases were defined as shown in Table 2 with phase 1 (−6.5 mm) corresponding to peak inhalation and phase 6 (+7.0 mm) corresponding to peak expiration.

**Table 2:** Phase definitions for Casper Phantom with Diaphragm Superior-Inferior (SI) Displacements.

| **Phase** | **Description** | **SI displacement (mm)** |
|---|---|---|
| 1 | Peak inhalation (Reference) | -6.5 |
| 2 | - | -3.5 |
| 3 | - | -0.5 |
| 4 | - | +2.5 |
| 5 | - | +5.5 |
| 6 | Peak Expiration | +7.0 |
| 7 | - | +4.0 |
| 8 | - | +1.0 |
| 9 | - | -2.0 |
| 10 | - | -5.0 |

Each 3D scan was generated from 2D X-ray projections at 397 imaging angles over one 200*^◦^* arc to produce a total of 3970 2D projections. The Casper images were also resized during training with 3D images resized to dimensions 128 x 128 x 128 and 2D projections resized to dimensions 128 x 128 due to memory constraints.

### 2.3 VoxelMap architecture

VoxelMap is a neural network which takes as input a target 2D projection, a reference 2D projection corresponding to phase 1, and a reference 3D image corresponding to phase 1 and outputs the predicted target 3D image corresponding to the target 2D projection. The VoxelMap architecture is composed of an encoder-decoder architecture followed by an integration layer and a spatial transformation module [19, 20]. Our proposed workflow for VoxelMap in interventional procedures is shown in Figure 3. Individual models for both phantoms were trained using the same VoxelMap architecture and training protocol.

**Figure 3:**
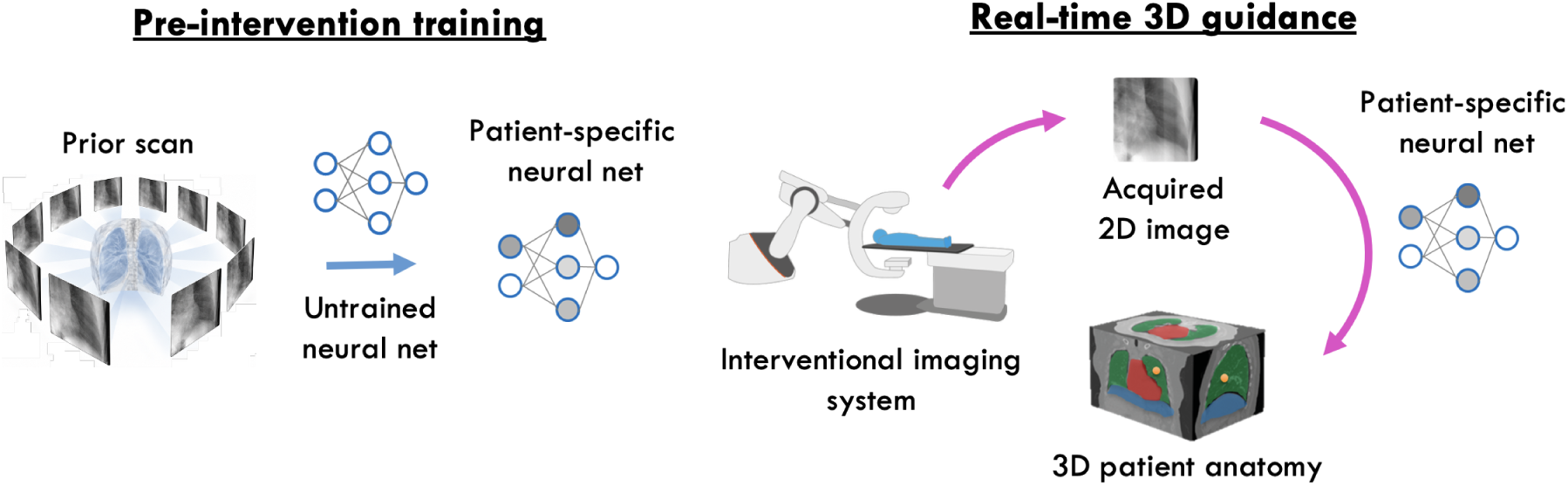
Our proposed workflow for utilizing VoxelMap to enable real-time 3D guidance in interventional procedures.

The objective function for training was the mean absolute error (MAE)

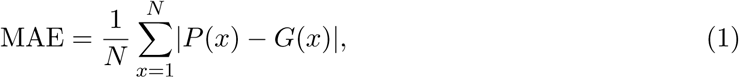

where for each voxel *x*, *P* is the value in the predicted 3D image and *G* is the value in the ground-truth 3D image.

We extend the VoxelMap architecture with a deep ensemble network. Following Lakshminarayanan (2016), 4 additional VoxelMap neural networks are instantiated with the same architecture and trained on the same training dataset and only differ by initial weights and data order, to produce a total of 5 models for uncertainty estimation [25]. The voxel-wise ensemble prediction is defined as the mean of the output of each neural network, and the voxel-wise ensemble uncertainty is defined as the standard deviation of the output of each neural network.

### 2.4 Experiments

Two deep ensembles were trained on each phantom. Each ensemble was trained with one phase excluded such that the training dataset for each ensemble consisted of all 2D projections that did not correspond to the excluded phase. The evaluation dataset consisted of the 397 2D projections corresponding to the excluded phase. The phases excluded for each phantom were (a) the phase corresponding to end exhalation as end exhalation is relatively anatomically stable and reproducible and (b) the phase corresponding to mid-inhalation to test whether more unstable imaging positions affect accuracy [28]. Specifically, phase 6 (AP 2.2 mm, SI 15.0 mm) and phase 8 (AP 0.6 mm, SI 3.6 mm) were excluded on the PixelPrint^4D^ phantom, and phase 6 (+7.0 mm) and phase 8 (−0.5 mm) were excluded on the Casper phantom. 2D projections in the training and evaluation dataset were linearly HU-normalized to [0, 1] using intensities from all projections in the training dataset.

The 5 neural networks within each ensemble were trained independently with the Adam learning algorithm with the learning rate 1 × 10^−5^ and batch size 4 for 100 epochs. The training dataset was randomly split 90/10 into training and validation sets. The model parameterisation with the lowest validation loss was saved for evaluation. Deep learning experiments used the PyTorch framework and were performed on a NVIDIA RTX 6000 Ada Generation GPU with 48 GB memory, requiring 6-8 hours for each neural network and 30-40 hours for each ensemble.

### 2.5 Evaluation metrics

Inference time is defined as the time taken for a single forward pass through each model in the ensemble. The mean and standard deviation of the model inference times was calculated over 392 2D projections corresponding to the excluded phase, excluding *n* = 5 warm-up projections.

Prediction accuracy of deformed 3D volumes compared to ground truth volumes was evaluated on deformations predicted from 2D projections of the respiratory phase excluded in the training of each ensemble. The similarity between predicted and ground truth volumes was quantified by the SSIM and peak signal-to-noise ratio (PSNR), all evaluated over the whole 3D volume.

To assess perceived structural accuracy, we used the SSIM:

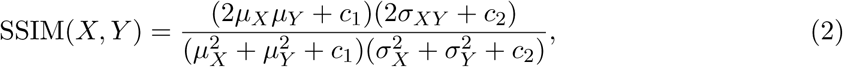

where *X* and *Y* are the ground-truth and predicted 3D images, respectively, and *c*_1_ and *c*_2_ are constants to stabilize division.

To assess the accuracy of the absolute intensities, we used the PSNR:

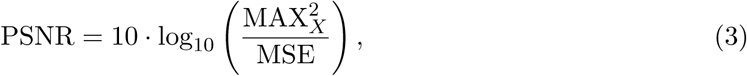

where MAX*_X_*is the maximum possible voxel intensity and MSE is the mean squared error.

The uncertainty-error correlation is defined as the Pearson correlation co-efficient between the MAE and ensemble uncertainty:

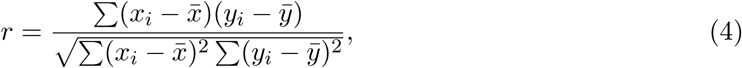

where *x* is the MAE of a predicted volume and *y* is the mean voxel-wise ensemble uncertainty of the same predicted volume.

## 3 Results

### 3.1 PixelPrint^4D^ experiments

#### 3.1.1 Evaluation of inference time

The mean and standard deviation of the inference time for each model was 10 *±* 4 ms in the phase 6 excluded ensemble and 9 *±* 3 ms in the phase 8 excluded ensemble, which is compatible with real-time processing.

#### 3.1.2 Evaluation of volumetric prediction accuracy

3D deformations predicted from the projection corresponding to the coronal view (projection 199) for all 8 phases for the phase 8 excluded ensemble are shown in Figure 4. The SSIM of one neural network trained with phase 8 excluded across 397 projection angles for all 8 phases is shown in Figure 5. Table 3 summarises the results of the prediction accuracy analysis. The SSIM and PSNR of the predicted volumetric constructions are compared with the ground-truth images in Figure 6.

**Figure 4:**
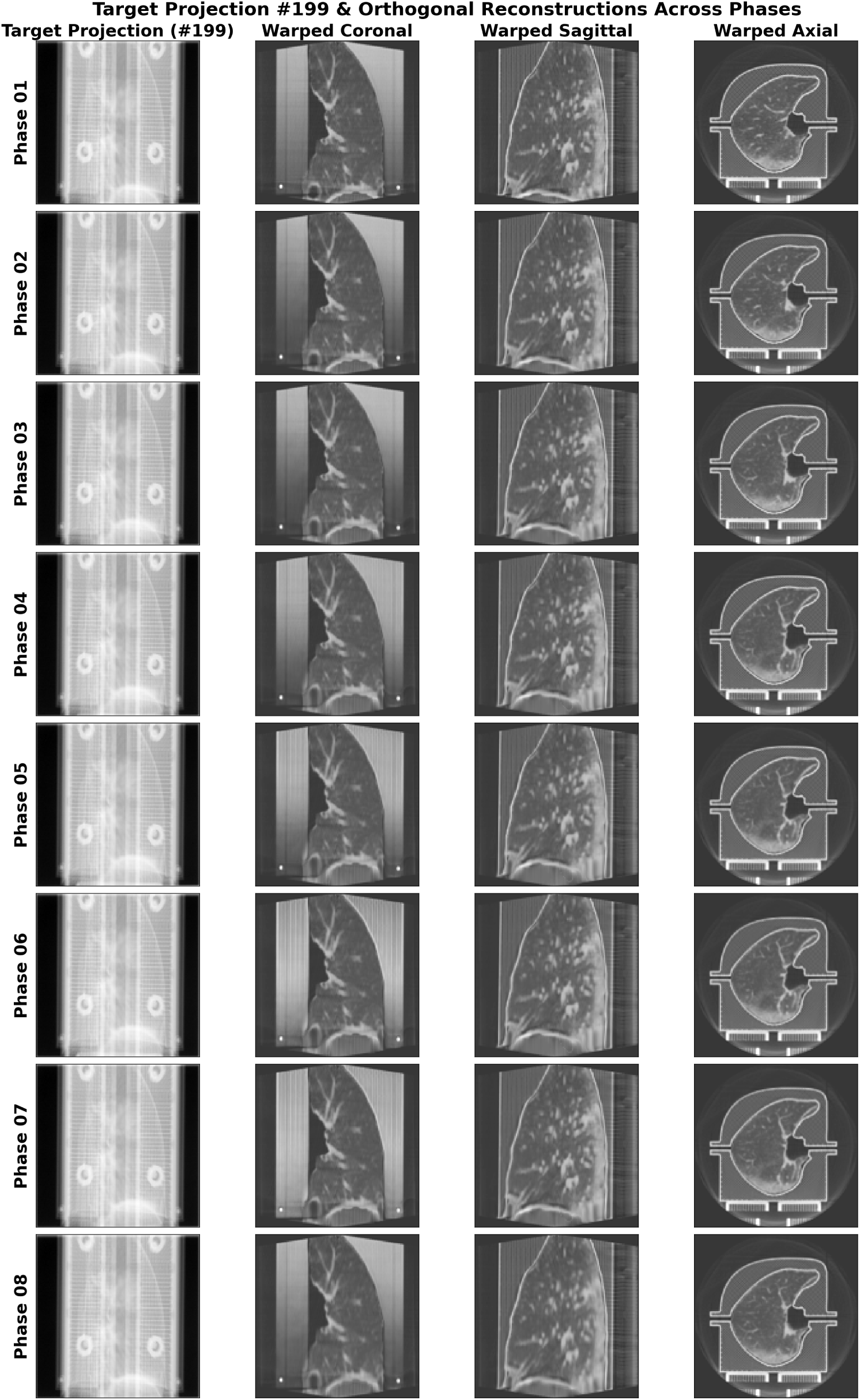
In each row, the target projection corresponding to a coronal view (#199) at each of the 8 phases and the corresponding mean predicted deformation by the PixelPrint^4D^ phase 8 excluded ensemble. From left to right: the target 2D projection (Target projection), then the corresponding mid-coronal (Warped Coronal), mid-sagittal (Warped Sagittal), and mid-axial (Warped Axial) projections of the predicted 3D deformation.

**Figure 5:**
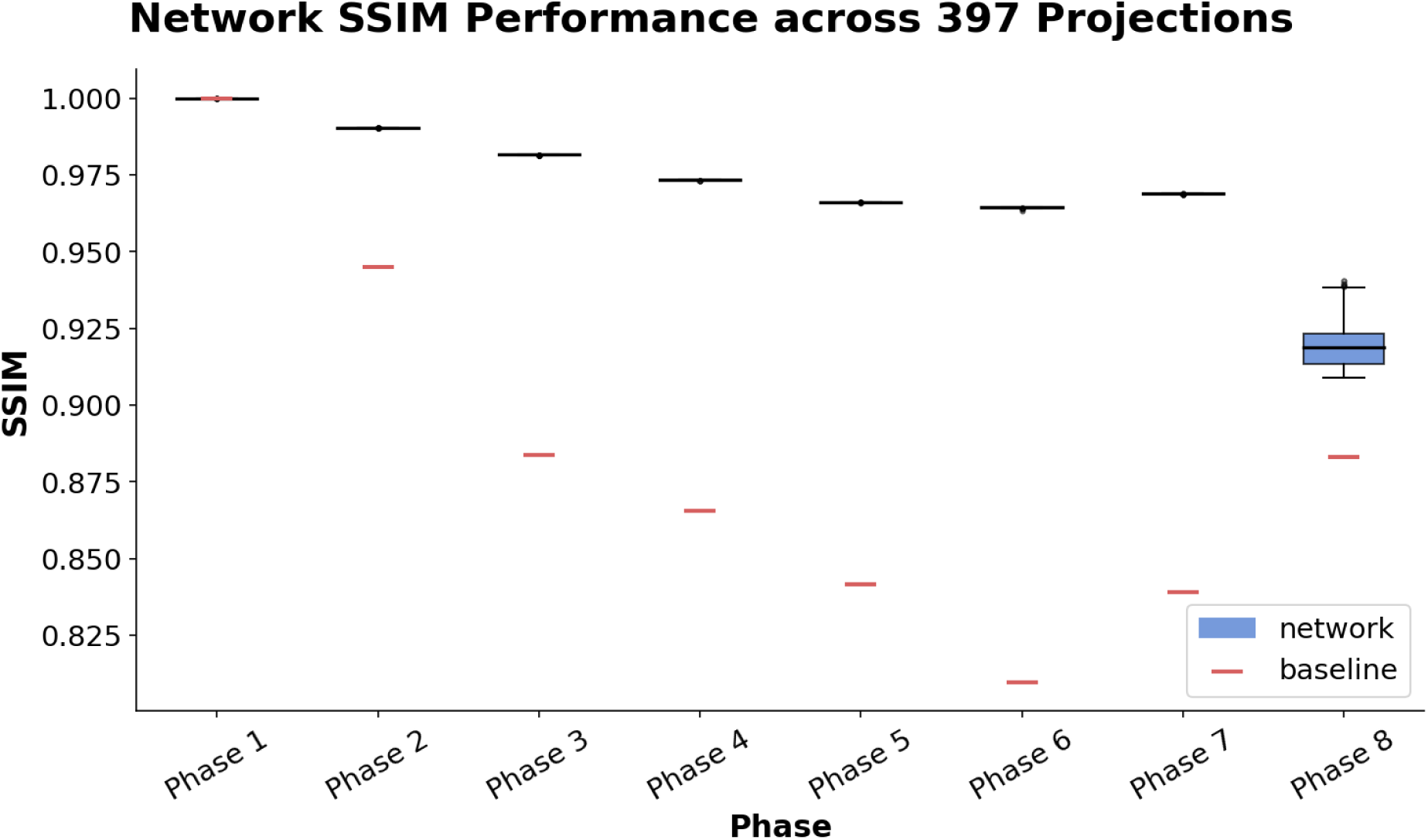
SSIM of the PixelPrint^4D^ phase 8 excluded network across 397 projection angles over all 8 phases compared to the reference phase 1 3D image (baseline).

**Figure 6:**
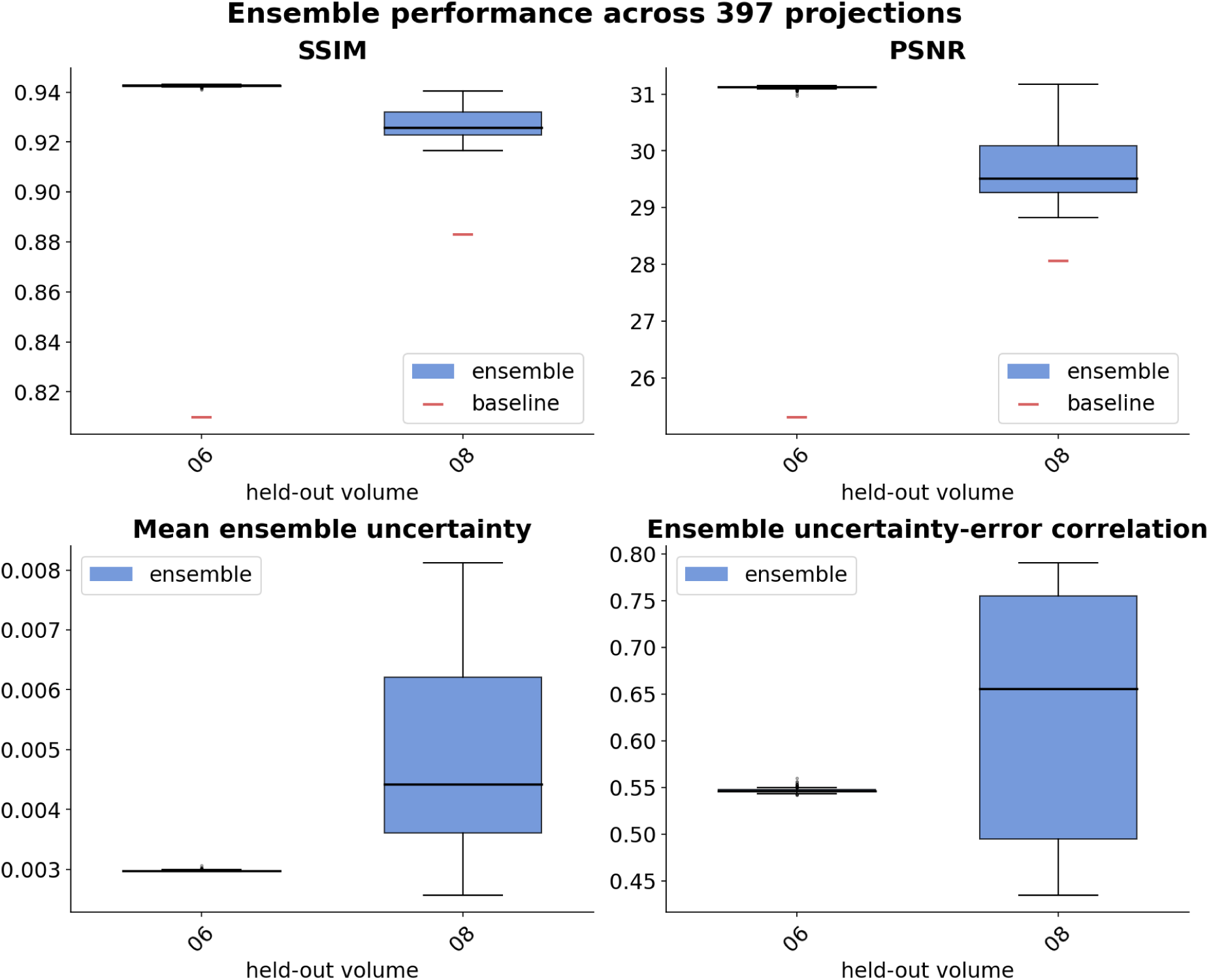
The SSIM, PSNR, mean ensemble uncertainty, and the uncertainty-error correlation of the predicted 3D image across the 397 2D projections in the PixelPrint^4D^ evaluation dataset for the phase 6 excluded and phase 8 excluded ensembles compared to the reference phase 1 3D image (baseline).

**Table 3:** Prediction accuracy analysis with Structural Similarity Index Measure (SSM), Peak Signal-to-Noise Ratio (PSNR), and Error-Uncertainty Correlation.

| **Phantom** | **Excluded phase** | **SSIM<sup>\*</sup>** | **PSNR<sup>\*</sup>** | **Error-Uncertainty Correlation<sup>\*</sup>** |
|---|---|---|---|---|
| PixelPrint<sup>4D</sup> | 6 | 0.9426 ± 0.0002 | 31.12 ± 0.02 | 0.547 ± 0.002 |
| PixelPrint<sup>4D</sup> | 8 | 0.9272 ± 0.0055 | 29.68 ± 0.52 | 0.628 ± 0.127 |
| Casper | 6 | 0.9689 ± 0.0004 | 41.39 ± 0.07 | 0.421 ± 0.018 |
| Casper | 8 | 0.9730 ± 0.0063 | 41.73 ± 1.11 | 0.651 ± 0.099 |

<sup>\*</sup> Values represent mean ± standard deviation.

VoxelMap demonstrated high prediction accuracy across both excluded respiratory phases. The phase 6-excluded ensemble achieved consistently higher and less variable SSIM than the phase 8-excluded ensemble (0.9426 *±* 0.0002 vs 0.9272 *±* 0.0055), with comparable differences observed in PSNR (Table 3). This trend is demonstrated across all projections in the evaluation dataset as shown in Figure 7. This was an improvement in SSIM relative to the reference phase 1 3D image (for the phase 6 excluded ensemble, an improvement of 0.1323 and for the phase 8 excluded ensemble, an improvement of 0.0443).

**Figure 7:**
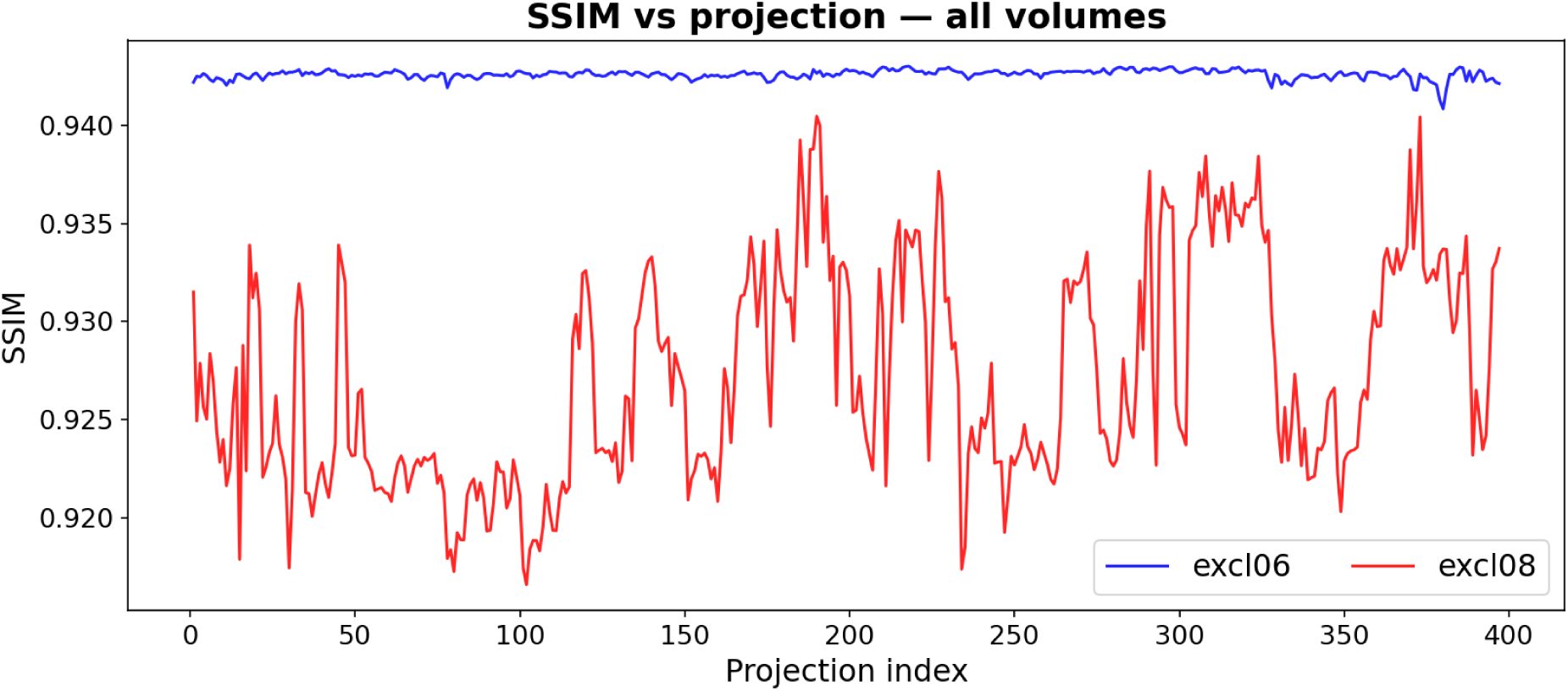
The SSIM of the deep ensemble across the 397 projections of the excluded phase for the phase 6 excluded and phase 8 excluded PixelPrint^4D^ ensembles.

#### 3.1.3 Evaluation of model uncertainty

The mean voxel-wise ensemble uncertainty for the predicted 3D images for each projection was consistently lower for the phase 6 excluded ensemble compared to the phase 8 excluded ensemble ((2.98 *±* 0.01) × 10^−3^ vs (4.84 *±* 1.51) × 10^−3^). The ensemble uncertainty over all projections in the evaluation dataset is shown in Figure 8.

**Figure 8:**
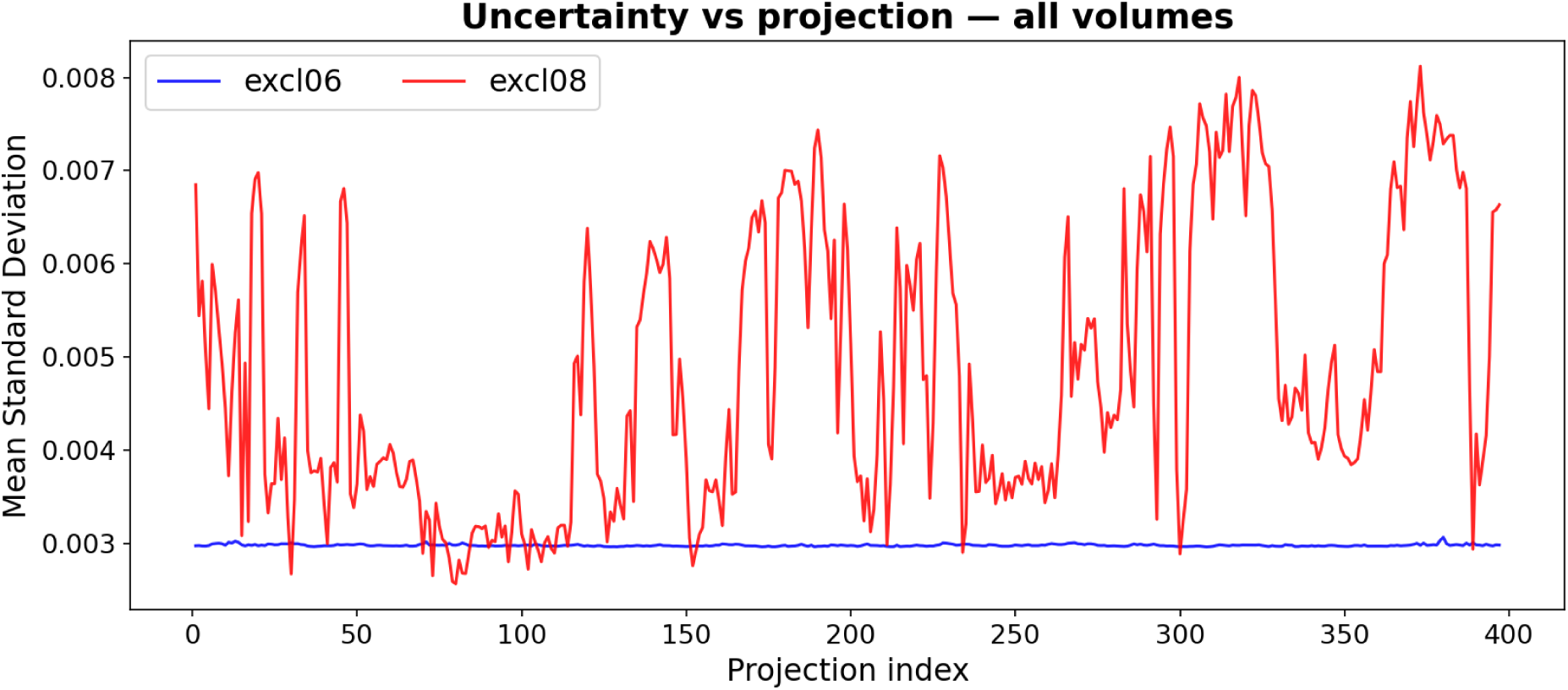
The mean ensemble uncertainty across the 397 projections of the excluded phase for the phase 6 excluded and phase 8 excluded PixelPrint^4D^ ensembles.

Ensemble uncertainty showed a positive correlation with prediction error, with Pearson correlation coefficients of 0.547 *±* 0.002 for phase 6 excluded and 0.628 *±* 0.127 for phase 8 excluded. Voxel-wise uncertainty maps demonstrated spatial correspondence between regions of increased uncertainty and increased prediction error. Uncertainty maps of the best and worst predicted 3D deformations by SSIM over both ensembles are shown in Figure 9.

**Figure 9:**
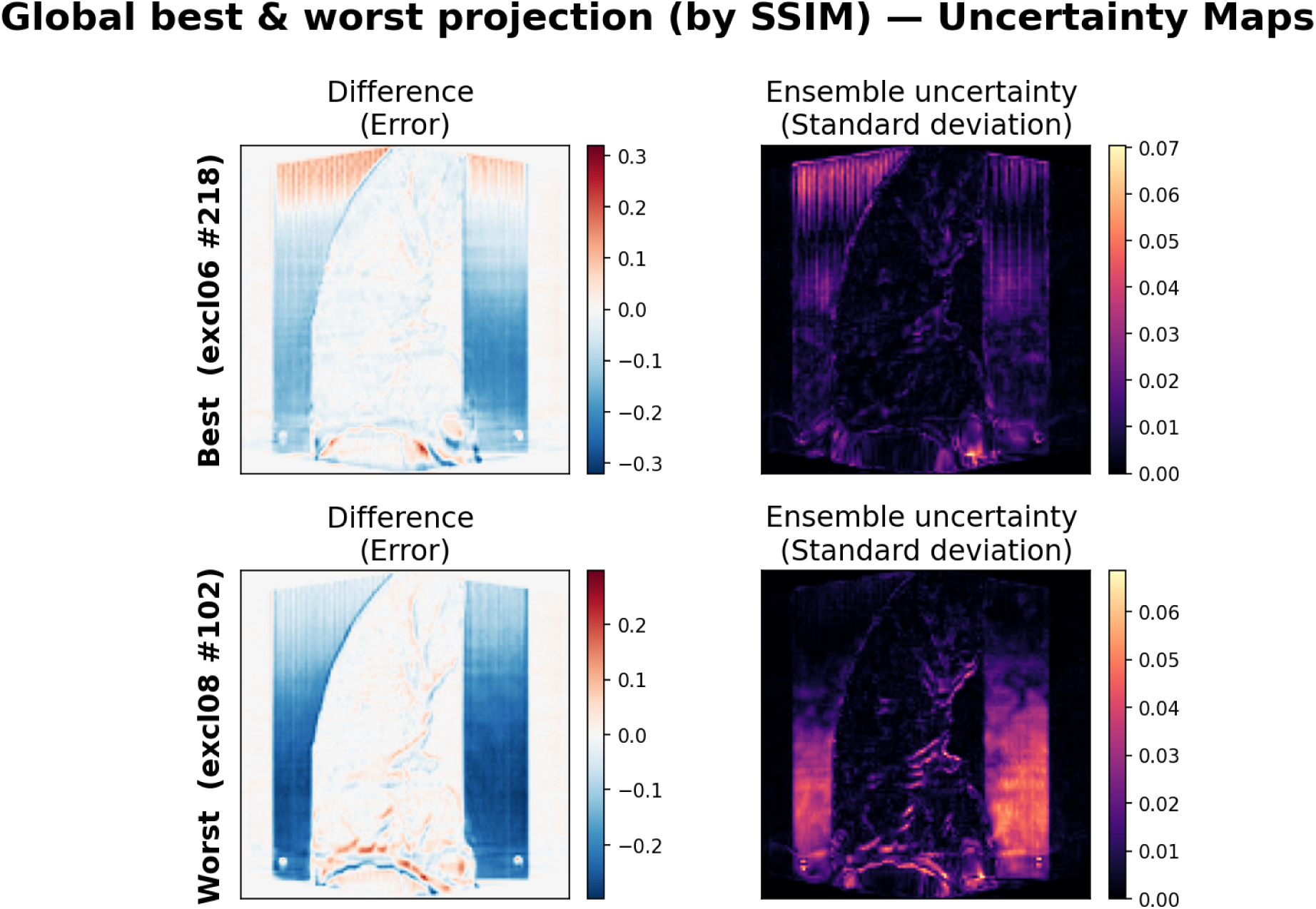
The best deformation (projection 218 in the phase 6 excluded ensemble) and the worst deformation (projection 102 in the phase 8 excluded ensemble) for the PixelPrint^4D^ ensembles are pictured in the top and bottom panel, respectively. The left panel is the differential map comparing predicted to ground truth 3D images and the right panel is the voxel-wise ensemble uncertainty.

### 3.2 Casper experiments

VoxelMap demonstrated similar performance when evaluated on projections of 3D images of the Casper thoracic phantom on a Siemens Artis Pheno. The mean and standard deviation of the inference time for each model was 9 *±* 3 ms in the phase 6 excluded ensemble and 10 *±* 4 ms in the phase 8 excluded ensemble. Prediction accuracy was consistently high across both excluded phases, with SSIM and PSNR values exceeding those achieved on PixelPrint^4D^ ensembles across all evaluated phases (Table 3). Deformations predicted from the projection corresponding to the coronal view (projection 199) for all 10 phases for the phase 8 excluded ensemble are shown in Figure 10. The SSIM of one neural network trained with phase 8 excluded across 397 projection angles for all 10 phases is shown in Figure 11.

**Figure 10:**
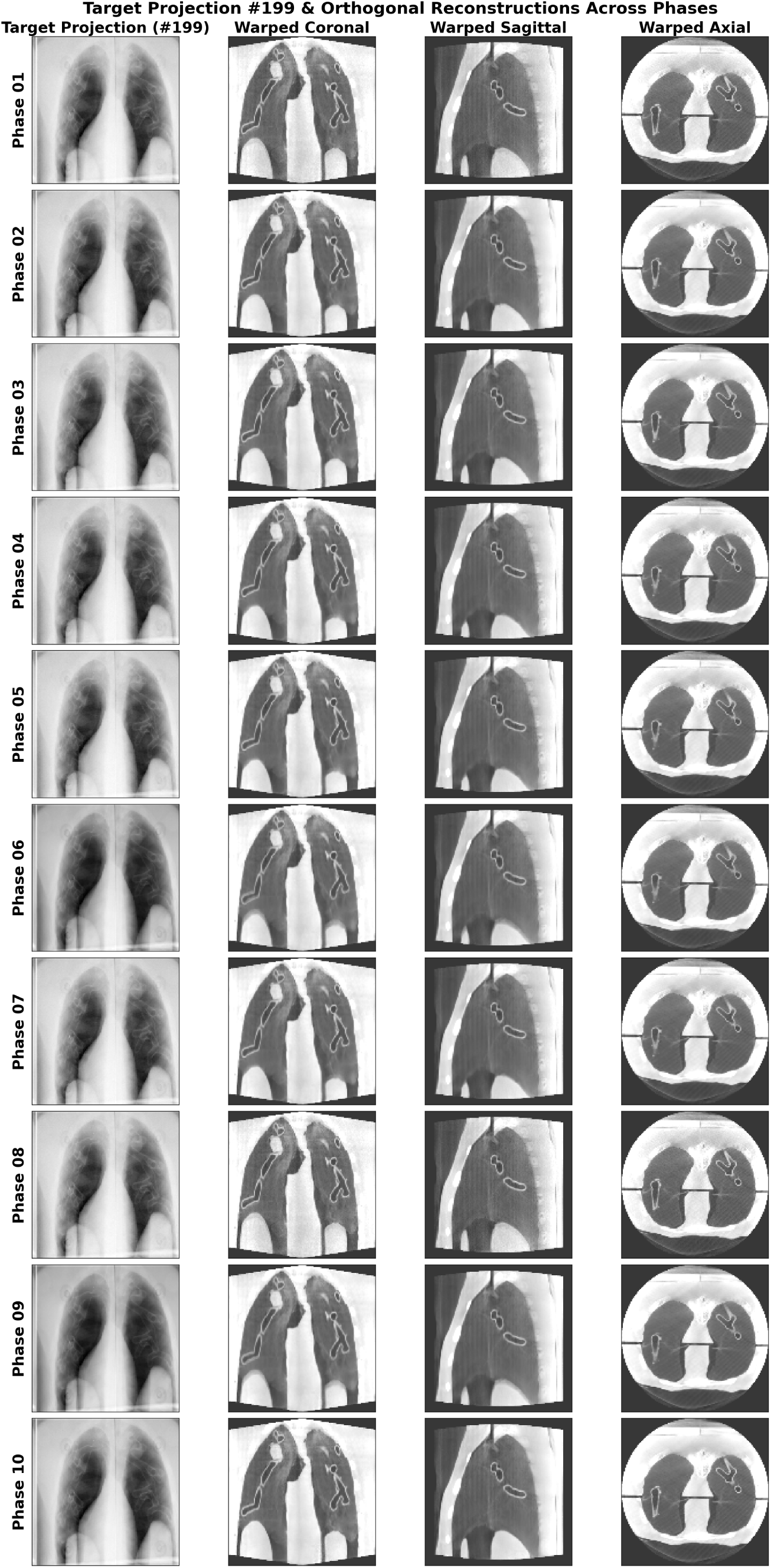
In each row, the target projection corresponding to a coronal view (#199) at each of the 10 phases and the corresponding mean predicted deformation by the Casper phase 8 excluded ensemble. From left to right: the target 2D projection (Target projection), then the corresponding mid-coronal (Warped Coronal), sagittal (Warped Sagittal), and mid-axial (Warped Axial) projections of the predicted 3D deformation.

**Figure 11:**
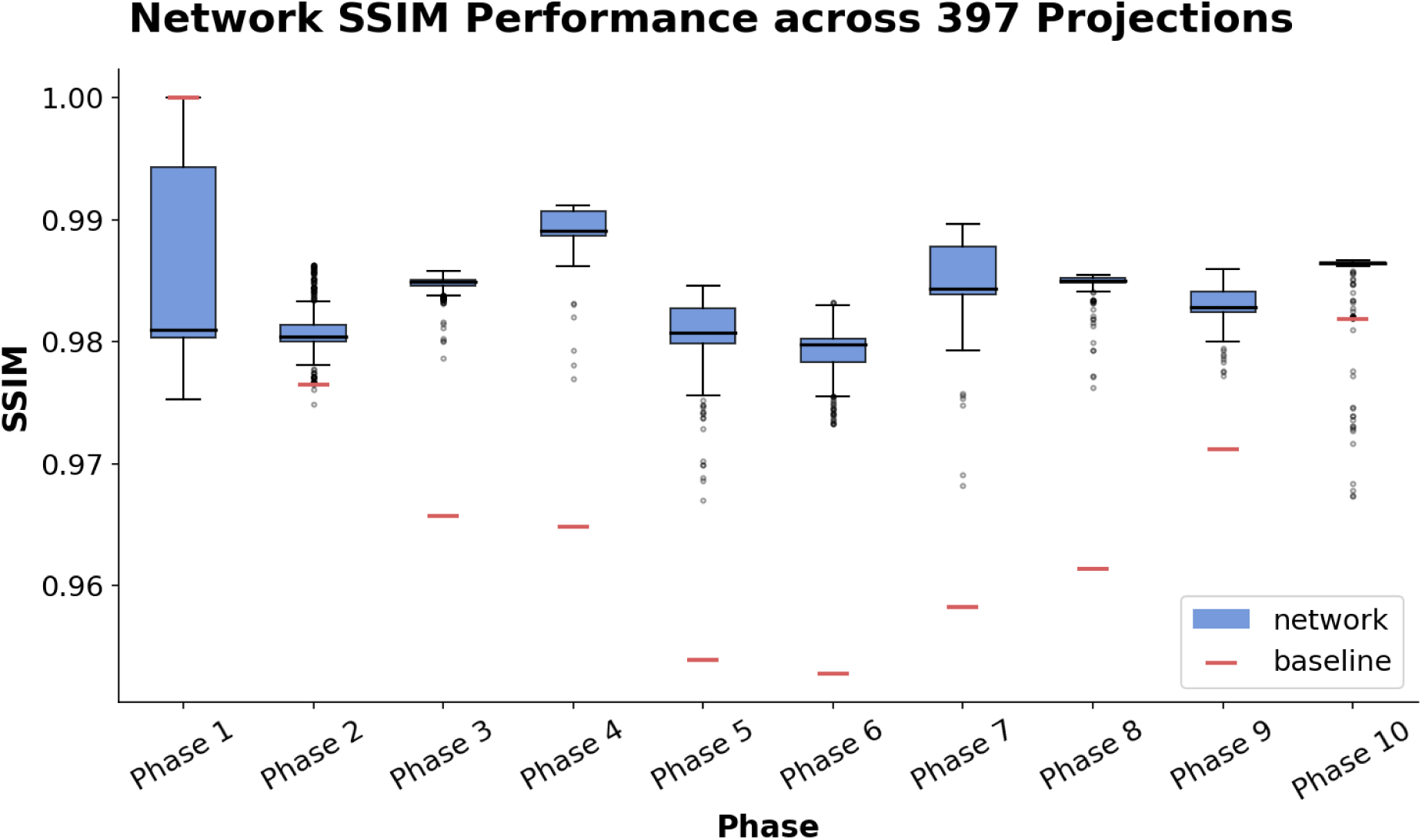
SSIM distribution of the Casper phase 8 excluded network across 397 projection angles over all 10 phases compared to the reference phase 1 3D image (baseline).

As observed with PixelPrint^4D^, the SSIM of the phase 6 excluded ensemble was less variable than that of the phase 8 excluded ensemble, presented in Figure 12. However, the SSIM of the Casper phase 6 excluded ensemble was lower than that of the phase 8 excluded ensemble (0.9689 *±* 0.0004 vs 0.9730 *±* 0.0063), as was the PSNR, presented in Figure 13.

**Figure 12:**
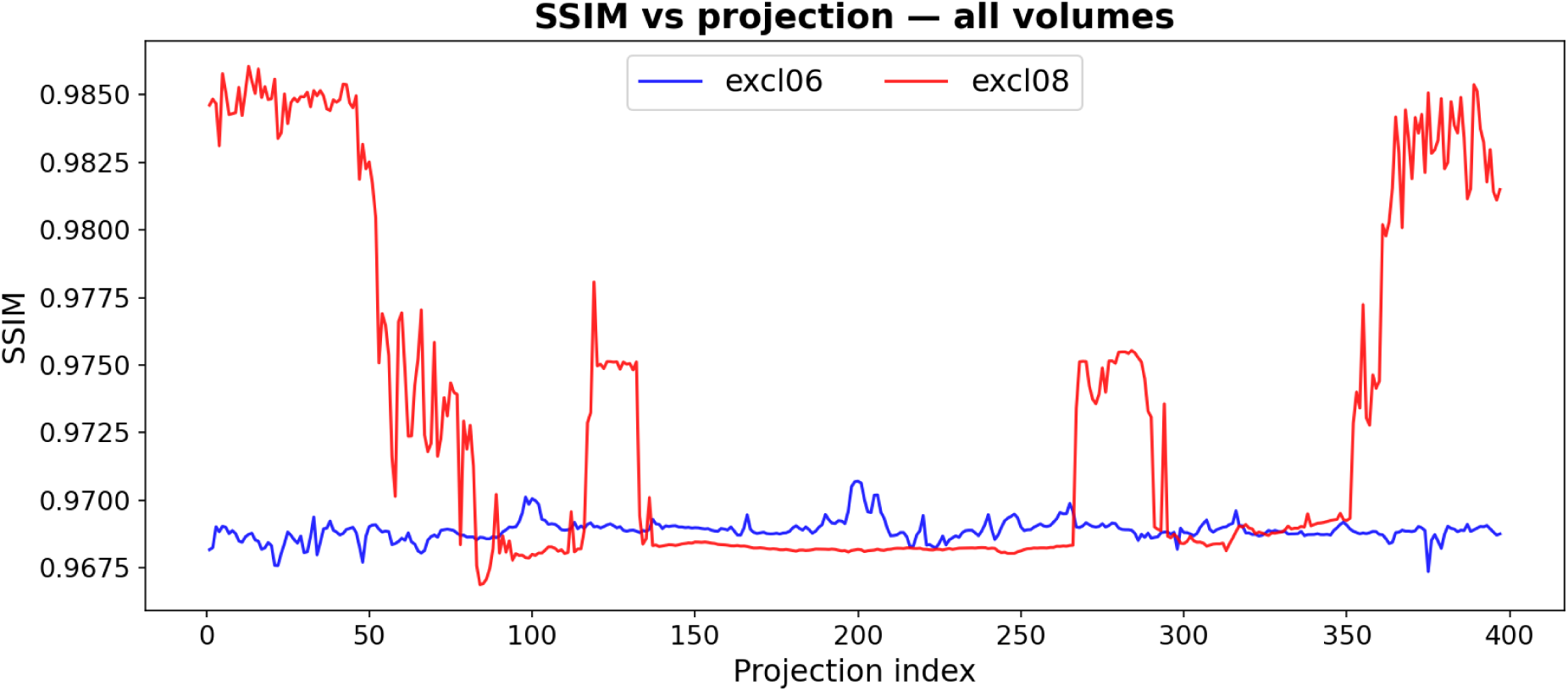
The SSIM of the deep ensemble across the 397 projections of the excluded phase for the phase 6 excluded and phase 8 excluded Casper ensembles.

**Figure 13:**
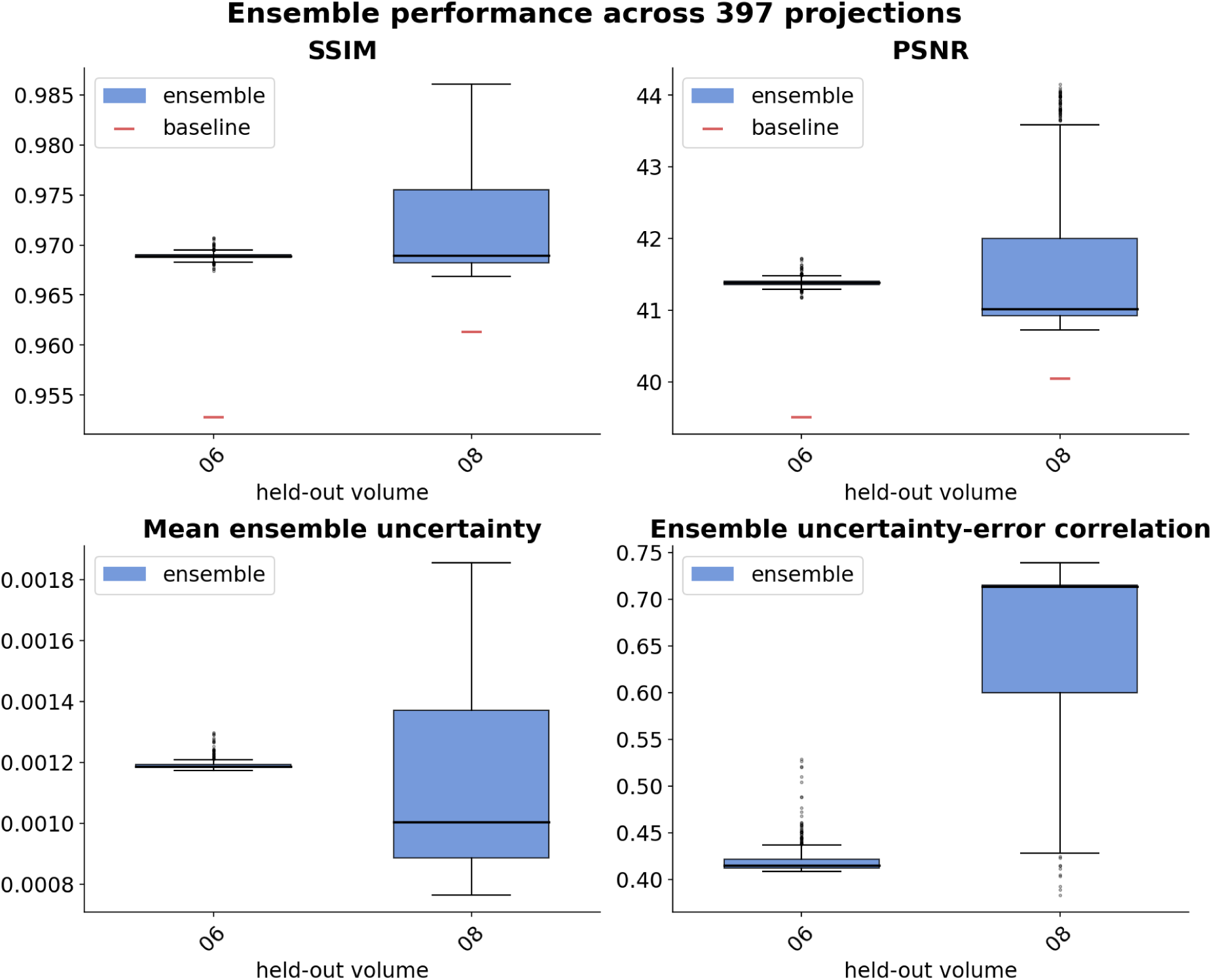
The SSIM, PSNR, mean ensemble uncertainty, and the uncertainty-error correlation of the predicted 3D image across the 397 2D projections in the Casper evaluation dataset for the phase 6 excluded and phase 8 excluded ensembles compared to the reference phase 1 3D image (baseline).

Similar to the PixelPrint^4D^ ensembles, the phase 6-withheld ensemble demonstrated a more consistent mean voxel-wise ensemble uncertainty than the phase 8-withheld ensemble ((1.19 *±* 0.02) × 10^−3^ vs (1.13 *±* 0.28) × 10^−3^), as pictured in Figure 14.

**Figure 14:**
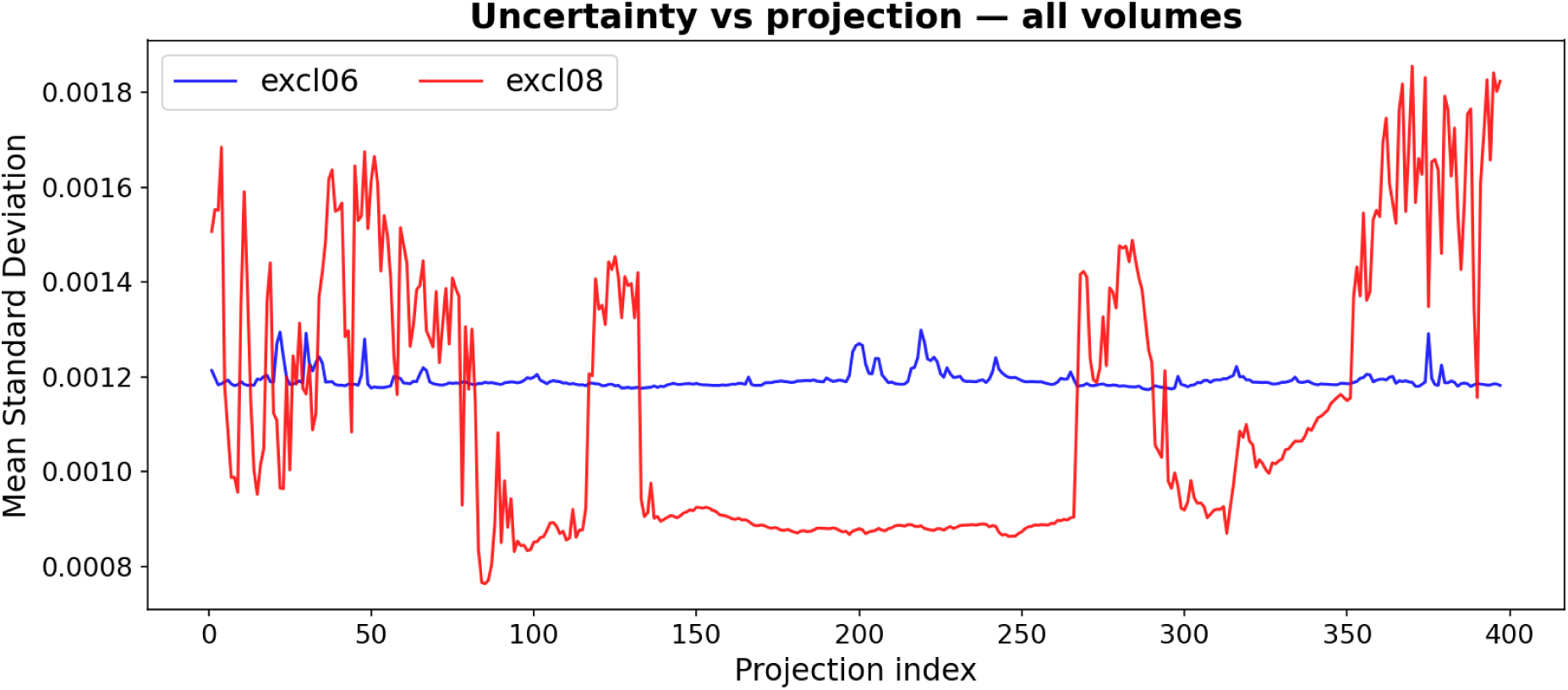
The mean ensemble uncertainty across the 397 projections of the excluded phase for the phase 6 excluded and phase 8 excluded Casper ensembles.

The error–uncertainty correlation were 0.421 *±* 0.018 for the phase 6 excluded ensemble and 0.651 *±* 0.099 for the phase 8 excluded ensemble, demonstrating a similar relationship between ensemble uncertainty and prediction error to that observed on the PixelPrint^4D^ ensembles. Uncertainty maps of the best and worst predicted 3D deformations by SSIM over both ensembles are shown in Figure 15.

**Figure 15:**
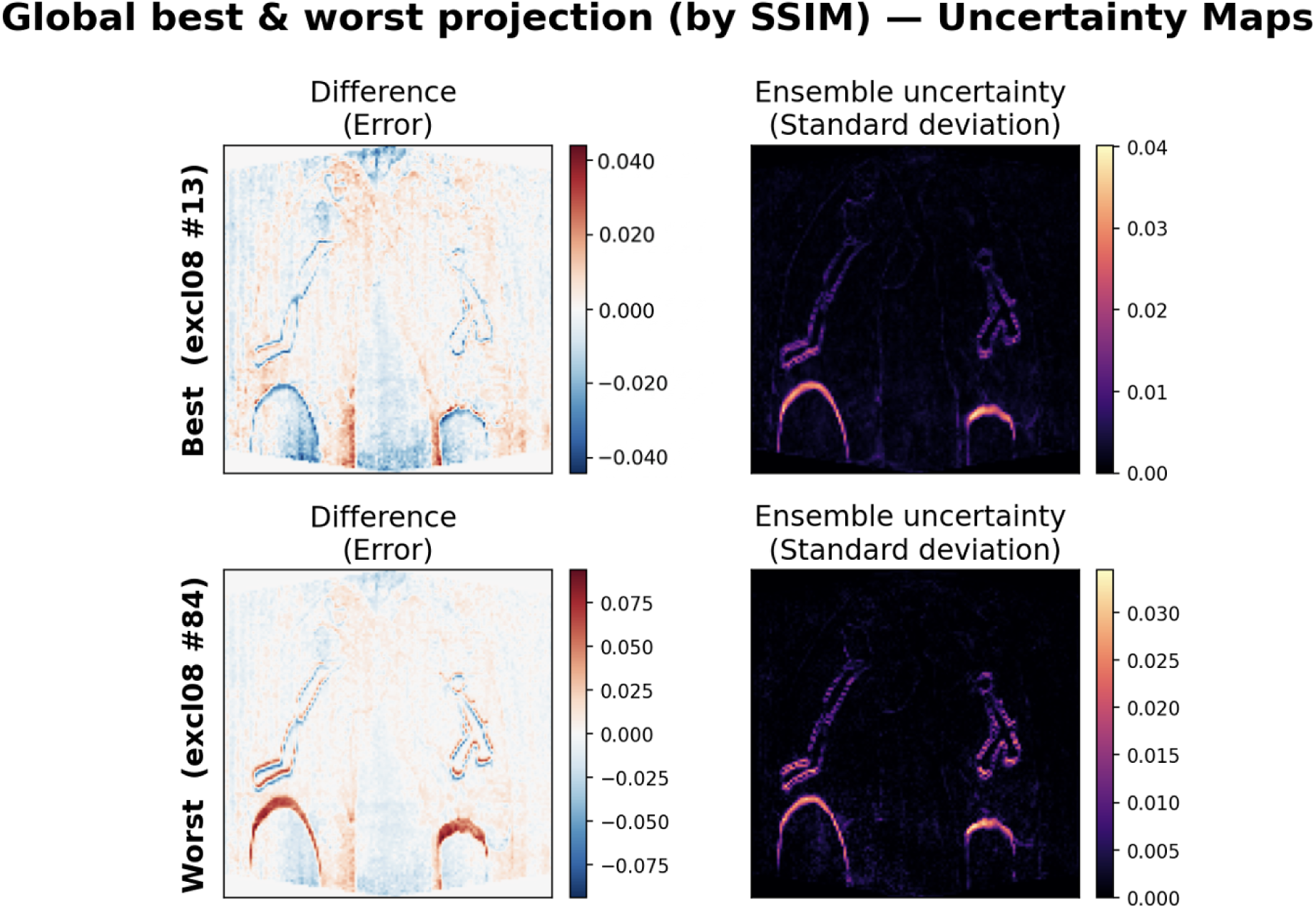
The best deformation (projection 13 in the phase 8 excluded ensemble) and the worst deformation (projection 84 in the phase 8 excluded ensemble) for the Casper ensembles are pictured in the top and bottom panel, respectively. The left panel is the differential map comparing predicted to ground truth 3D images and the right panel is the voxel-wise ensemble uncertainty.

## 4 Discussion

In this study, VoxelMap demonstrated accurate volumetric representations of respiratory anatomy from 2D projections acquired on a clinical interventional C-arm system. VoxelMap maintained high prediction accuracy for withheld respiratory phases across two physical respiratory phantoms while achieving sub-15 ms model inference times. These findings address key limitations of current interventional image-guidance techniques: fluoroscopy provides continuous procedural imaging but limited 3D anatomical information, whereas volumetric techniques including CBCT and DTS provide 3D information but require dedicated acquisitions and therefore do not provide continuously updated volumetric information throughout an intervention [13, 14, 17]. By deriving volumetric information from fluoroscopic projections, VoxelMap has the potential to complement existing interventional workflows.

Image-guided navigation is increasingly used in interventional procedures to localize anatomical targets and provide navigational guidance [29, 30]. However, the spatial relationship between targets, anatomy, and instruments can change during an intervention [15, 14]. For thoracic procedures, pre-procedural CT can become inaccurate as patient anatomy changes with respiratory motion [31, 32, 33]. Intra-procedural CBCT addresses some of these limitations by providing intermittent real-time volumetric information, but requires a 200*^◦^* rotational acquisition and anatomical accuracy may be reduced by respiratory motion during image acquisition [13, 34, 35]. Therefore, there is an unmet need for approaches that can provide volumetric information without interrupting the continuous fluoroscopic workflow.

Although this study focuses on thoracic interventions, potential applications of our approach extend across image-guided interventions which require 3D localisation of anatomy and instruments using 2D continuously acquired fluoroscopic images. Intra-procedural respiratory and other forms of anatomical motion make maintaining correspondence between 2D projections and the underlying 3D anatomy challenging [15, 36]. Therefore, a method to produce volumetric information directly from continuous fluoroscopic data may reduce this difficulty.

To our knowledge, our study represents the first demonstration of volumetric imaging of respiratory anatomy potentially suitable for continuous real-time volumetric imaging in an interventional setting. VoxelMap achieved high structural similarity across withheld respiratory phases in both physical phantoms, with SSIM values exceeding 0.92 in all experiments (Table 3). In addition, model inference times were less than 15 ms across both phantoms, demonstrating computational latency likely compatible with continuous real-time processing of fluoroscopic images. Importantly, these results also demonstrate the first validation of VoxelMap using projections acquired on a clinical fixed-room C-arm angiography system.

In addition, our paper represents the first validation of VoxelMap on the Pixelprint^4D^ phantom and the Casper phantom. Evaluation across both respiratory phantoms demonstrates that VoxelMap can generalise across distinct physical respiratory phantoms. VoxelMap achieved high SSIM values across both phantoms, with higher SSIM than the baseline reference in both phantoms. Prediction performance was higher on Casper than PixelPrint^4D^, with SSIM values of 0.969–0.973 compared with 0.927–0.943, respectively. This difference may reflect differences in respiratory deformation and phantom characteristics rather than an inherent difference in model performance. Across projection angles, the variation in SSIM for the Casper phase 8 excluded ensemble was less than 0.02, indicating relatively consistent reconstruction performance across acquisition angles. Overall, the consistently high prediction accuracy across both phantoms demonstrates the reproducibility of our approach across different physical respiratory models.

The potential use of AI to produce predicted volume deformations means that the ability to identify areas where the model may be unreliable is important for clinical adoption [21, 22, 23]. In both phantoms, ensemble uncertainty was positively correlated with prediction error with correlation coefficients ranging from 0.42-0.65, suggesting that uncertainty may be an informative indicator of deformation reliability. The observed correlation indicates that ensemble uncertainty may be a useful indicator of prediction reliability [37]. We also noted more stable uncertainty across projections on peak exhale-withheld ensembles (phase 6 excluded on PixelPrint^4D^ and Casper) compared to mid-inhale-withheld ensembles (phase 8 excluded on PixelPrint^4D^ and Casper). This may reflect the relatively stable anatomy at peak expiration [28].

The 4D CBCT images used as training data and volumetric ground truth do not represent routine interventional imaging. However, our primary aim was to evaluate the volumetric information generated by VoxelMap from projections acquired on a clinical interventional C-arm, while minimizing other changes from the previously published VoxelMap framework, which used 4D CT [19, 20]. The use of 4D CBCT therefore enabled the evaluation of projections acquired on a clinical interventional system while providing volumetric ground truth for quantitative assessment of prediction accuracy. In particular, the availability of volumetric ground truth across two physical thoracic respiratory phantoms provides a rigorous basis for evaluating the prediction of dynamic 3D anatomy. Further work is required to investigate network training strategies using data that more closely reflect clinically available imaging.

There are several limitations. First, as a phantom study, we do not capture effects of changes in patient respiratory motion between pre-procedural imaging and interventional procedures. Further validation of VoxelMap on patient CT images using realistic imaging protocols is required. Second, two physical phantoms do not capture the variability in anatomy and respiratory motion across patient populations. Additional evaluation of VoxelMap across diverse respiratory motion patterns and patient datasets is required to establish generalisability. Third, although we have demonstrated that VoxelMap inference times are consistent with use in a real-time interventional workflow, we did not evaluate the latency of the complete acquisition to visualisation pipeline. Further validation of VoxelMap in a more complete interventional workflow is warranted. Fourth, we have not established the clinical usefulness of the model error-uncertainty correlation. Further validation using patient data and clinically meaningful error thresholds is required to determine whether uncertainty can reliably identify clinically unacceptable reconstructions.

## 5 Conclusion

In conclusion, this study demonstrated that VoxelMap can generate accurate volumetric deformations of respiratory anatomy from 2D projections acquired on a clinical fixed-room C-arm angiography system, with sub-15 ms model inference times across two physical respiratory phantoms. This represents the first application of VoxelMap to projections acquired on a clinical fixed-room C-arm angiography system and the first validation on the physical PixelPrint^4D^ and Casper respiratory phantoms. We also establish the ensemble uncertainty as a potentially useful indicator of prediction reliability. To our knowledge, this is the first demonstration of a volumetric imaging technique which has the potential to provide continuously updated volumetric imaging during an intervention.

## Data Availability

The data that support the findings of this study are available from Siemens Healthineers but restrictions apply to the availability of these data, which were used under license for the current study, and so are not publicly available. Data are however available from the authors upon reasonable request and with permission of Siemens Healthineers.

https://github.com/Image-X-Institute/Voxelmap

## Acknowledgements

The authors acknowledge the facilities as well as the scientific and technical assistance of Sydney Imaging Core Research Facility

